# Tracking the Dynamic Trajectories: A Global-to-Local Pharmacovigilance Analysis of GLP-1 Receptor Agonists

**DOI:** 10.64898/2026.05.28.26354401

**Authors:** Shuyu Lu, Xiaoyang Ruan, Liwei Wang, Xiaomeng Wang, Murali Sameer, Hongfang Liu

**Affiliations:** McWilliams School of Biomedical Informatics, the University of Texas Health Science Center at Houston; Obesity Medicine, Memorial Hermann Health System

## Abstract

Although GLP-1/GIP receptor agonists demonstrate unprecedented weight-loss efficacy, their rapid clinical adoption has revealed significant real-world tolerability challenges. To evaluate their dynamic safety profiles, we developed a macro-to-micro pharmacovigilance framework by combining global FAERS reports with local UT-Physician EHR. Macroscopically, we distilled 17 shared adverse events across the drug class from FAERS with disproportionality analysis. Microscopically, local EHR data (289,655 longitudinal treatment sessions across 71,316 patients) revealed 51.6% of GLP-1 sessions terminated within 90 days. Furthermore, temporal-stratified logistic regression demonstrated that initial exposure (0–30 days) correlated strongly with nausea and vomiting, which attenuated in extended sessions, whereas extended exposure (>2 years) uncovered late-onset risks, notably incident hepatic steatosis. Ultimately, this time-aware framework reveals that GLP-1 safety profiles are profoundly duration-dependent, providing critical insights into both acute intolerances and long-term medication safety.

## Introduction

Obesity has long been recognized as a significant threat to global public health, currently affecting over a third of the world’s population ^1,2^. It markedly increases the risk of chronic comorbidities, including type 2 diabetes, cardiovascular disease, and certain cancers, while imposing a substantial socioeconomic burden worldwide ^3,4^. Consequently, substantial efforts have been dedicated to combatting this epidemic. During the past decades, a wide variety of therapies have emerged for obesity treatment, such as bariatric surgeries, anti-obesity medicines (AOMs), hydrogels, vagal nerve blockade, along with traditional dietary control and exercise. However, no single or combination of therapies performs consistently across individual patients, either in the short or long term ^5–8^.

Recently, the therapeutic landscape has been transformed by Glucagon-like peptide-1 (GLP-1) and glucose-dependent insulinotropic polypeptide (GIP) receptor agonists (RAs). As the fastest-growing segment of the pharmaceutical market, these therapies have demonstrated unprecedented efficacy in weight reduction. However, their rapid clinical adoption has outpaced our comprehensive understanding of their real-world safety profiles.

Despite their therapeutic potential, GLP-1/GIP RAs exhibit significant interindividual variability in tolerability ^9–11^. While some patients achieve meaningful weight loss, a substantial subset suffers from severe adverse events (ADEs). These side effects are not merely transient nuisances but revealed in a recent study as the leading cause of treatment discontinuation, second only to insurance-related issues ^12^. To date, our understanding of these specific adverse events primarily stems from randomized controlled trials (RCTs) and subsequent meta-analyses. Generally, most clinical trials meticulously track and report side effects, along with their severity, throughout predefined follow-up periods. Both mild gastrointestinal ADEs like nausea and vomiting, and severe conditions such as pancreatitis ^13–15^, cholelithiasis ^16^, and sinus tachycardia ^17–19^ are reported. Despite establishing a crucial safety baseline, these traditional trial methodologies possess inherent limitations. They typically enforce strict enrollment criteria, rely on limited sample sizes for rare events, and are constrained by relatively short follow-up durations. Consequently, they often fail to capture the long-term, dynamic trajectory of adverse events (ADEs) in diverse real-world populations.

To overcome these limitations, existing pharmacovigilance primarily relies on spontaneous reporting systems like the FDA Adverse Event Reporting System (FAERS), which offers a global scale for signal detection but inherently lacks granular clinical and temporal context. Conversely, local Electronic Health Records (EHR) provide rich, patient-level longitudinal data. While some previous studies have integrated EHRs into pharmacovigilance ^20,21^, they primarily focused on static signal validation, largely underutilizing the continuous temporal data required to track long-term medication trajectories. This temporal gap is particularly critical because obesity and its related chronic conditions demand sustained pharmacological interventions; therefore, it is imperative to study how AOM-related ADEs dynamically evolve from short-term intolerances to long-term risks.

Therefore, to overcome the inherent limitations of static signal detection, we propose a macro-to-micro sequential pharmacovigilance framework to systematically evaluate the safety profiles of GLP-1/GIP RAs against established AOMs. Functioning as a data-driven funnel, we first leverage global FAERS reports to capture a comprehensive spectrum of baseline ADE signals. We then filter and translate these signals into a large-scale local EHR system for strict clinical validation. With established longitudinal treatment sessions identified from local EHR, we transition from static signal detection to dynamic temporal analysis. Ultimately, this work elucidates treatment-duration-dependent toxicity, generating actionable hypotheses for safer, more personalized obesity management.

## Methods

### Study Design and Framework

This study employed a retrospective, two-phase observational design to evaluate the dynamic safety profiles of GLP-1/GIP RAs compared to baseline AOMs (**Figure 1**). The first phase (macro-level) consisted of a large-scale disproportionality analysis using the FDA Adverse Event Reporting System (FAERS) to identify and refine a core set of shared adverse event (ADE) signals. The second phase (micro-level) involved targeted clinical validation within the longitudinal UT Physician EHR cohort. During this phase, the core FAERS signals were semantically mapped to the local database, where patient records were structured into continuous treatment sessions. Finally, by applying temporal-stratified multivariate logistic regression models to these sessions, we quantitatively characterized how the risks of specific ADEs evolve across varying treatment durations.

**Figure 1.**
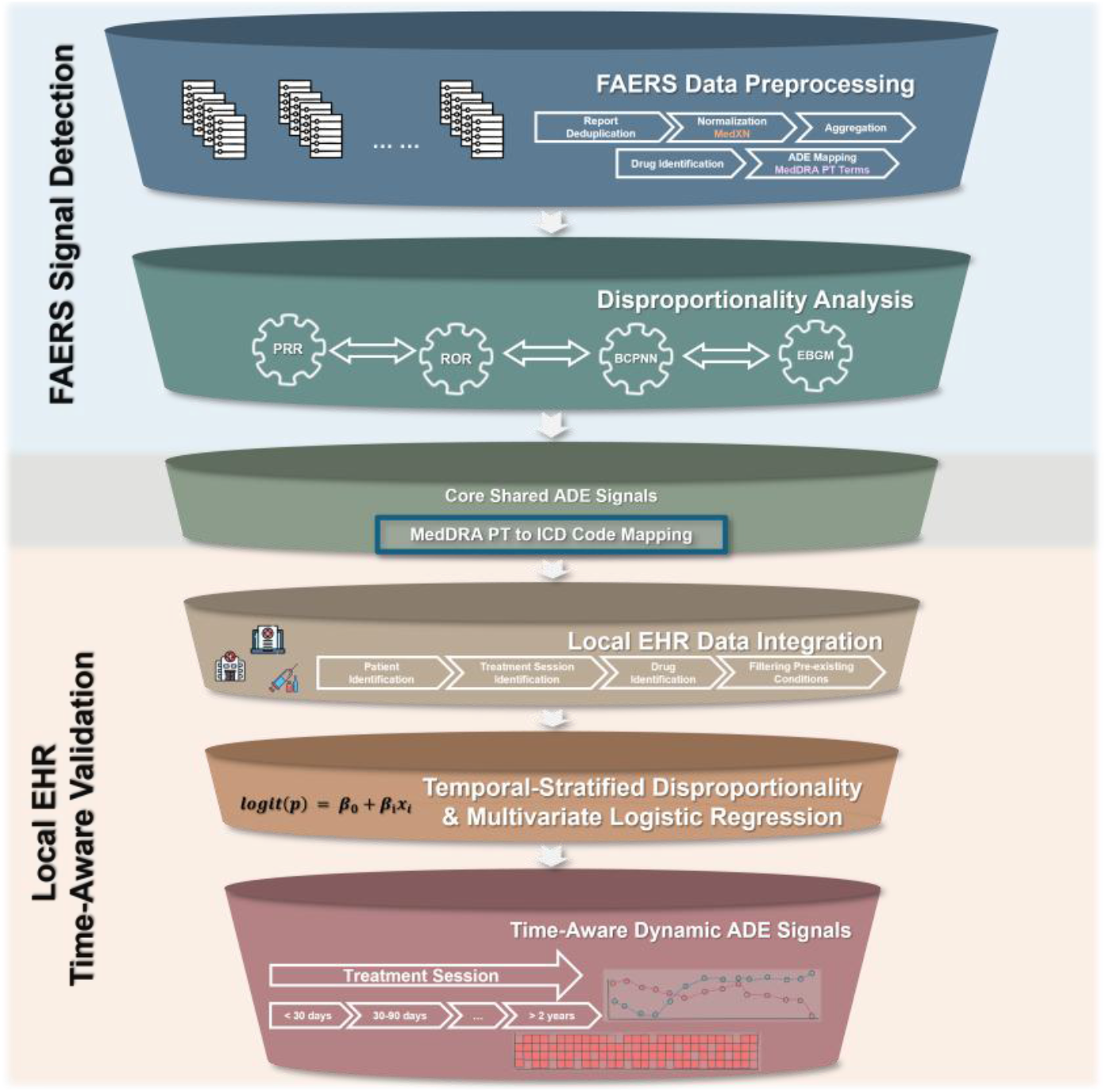
A Macro-to-Micro Sequential Pharmacovigilance Study Framework

### Anti-Obesity Medications (AOMs) Target and Baseline

To comprehensively investigate the adverse event spectrum of GLP-1/GIP RAs, we identified 20 medications commonly utilized for weight management, classifying them into four functional categories: (1) GLP-1/GIP RAs (study targets); (2) Core AOMs; (3) Weight-affecting Metabolics; and (4) Background medications often co-prescribed for weight control. This AOM list represents a robust, broad baseline of established metabolic treatments, enabling a comprehensive comparative safety analysis of the newer GLP-1/GIP therapies. The complete drug list and pharmacological classes are detailed in **Table 1**.

**Table 1:** Included Anti-Obesity Medications.

| Table 1: Included Anti-Obesity Medications |  |  |
| --- | --- | --- |
| Category | Included Drugs (Ingredient Level) | Pharmacological Class |
| Study Targets | Semaglutide, Liraglutide, Tirzepatide | GLP-1 / GIP receptor agonists |
| Core AOMs | Phentermine, Bupropion†, Naltrexone†, Orlistat, Topiramate, Zonisamide | Sympathomimetic, NDRI, Opioid receptor antagonist, Pancreatic lipase inhibitor, GABA/Sodium Modulation |
| Metabolics | Metformin, SGLT2 inhibitors (e.g., Dapagliflozin, Empagliflozin), Pioglitazone | Biguanide, SGLT2 inhibitors, PPAR-γ agonist |
| Background Meds | Lisdexamfetamine, Saxagliptin, Linagliptin | Stimulant, DPP-4 inhibitors |

† Often co-prescribed for weight management.

### FAERS dataset and Preprocessing

Serving as the broad entry point of our data-driven funnel, the FAERS was utilized for large-scale, global signal detection. Because FAERS contains spontaneous reports with redundant entries and arbitrary drug names (including trade names, abbreviations, and typographical errors) ^22^, we adopted a validated informatics preprocessing pipeline ^23^ consisting of three critical stages: de-duplication, normalization, and data aggregation.

First, redundant reports were removed using *primaryid* and *caseid* to ensure that only the most recent version of each case was retained. Second, normalization was performed in two steps: (1) raw drug entries were processed via MedXN ^24^ to map medication names and administration details to standardized RxNorm codes; and (2) adverse event terms were mapped to the Medical Dictionary for Regulatory Activities (MedDRA, v28.0) Preferred Terms (PTs) adopting from the Unified Medical Language System (UMLS) Metathesaurus ^25^. Finally, during data aggregation, records were synthesized at the active ingredient level (e.g., mapping both Ozempic and Wegovy to Semaglutide) to facilitate class-wide safety assessments. This pipeline was applied to the FAERS Quarterly Data Extract Files spanning from 2014 Q1 to 2025 Q4, establishing the foundational dataset for subsequent macro-level signal detection.

### Disproportionality Analysis

To identify comprehensive Signals of Disproportionate Reporting (SDRs) ^26^ from the FAERS dataset, we employed four widely validated pharmacovigilance algorithms based on standard two-by-two contingency tables. These included two frequentist methods: the Proportional Reporting Ratio (PRR) ^27^ and the Reporting Odds Ratio (ROR) ^28^; as well as two Bayesian shrinkage methods designed to minimize false positives in sparse data: the Bayesian Confidence Propagation Neural Network (BCPNN) ^29^ and the Multi-item Gamma-Poisson Shrinker (MGPS) ^30^.

A drug-ADE association was defined as a positive safety signal if it met the established statistical thresholds: (1) for PRR and ROR, the lower bound of the 95% confidence interval (CI) > 1 with a minimum case count of 3 ^31^; (2) for BCPNN, the Information Component (IC) lower bound (IC_025_) > 0 ^31^. The IC measures whether an adverse event is reported alongside a specific drug significantly more often than would be expected by random chance across the entire database. The “IC_025_” represents the lower bound of its 95% confidence interval. Therefore, a value strictly greater than zero provides strong statistical confidence that the association is a genuine safety signal; and (3) for MGPS, the Empirical Bayes Geometric Mean (EBGM) lower bound (EB_05_) > 2 ^32^. By intersecting the signals detected across all target GLP-1/GIP RAs, we distilled a core set of shared ADEs for subsequent clinical verification.

### MedDRA to ICD Mapping via UMLS

Since FAERS encodes adverse events in MedDRA while the local EHR utilizes ICD codes, we utilized UMLS Metathesaurus ^33^ 2025 AB version ^34^ to perform semantic cross-walking, translate the core signals discovered in FAERS into target conditions for local EHR validation. The mapping was conducted by linking MedDRA Preferred Terms (PTs) to Concept Unique Identifiers (CUIs) via the MRCONSO table (retaining only active, non-suppressed terms), followed by a CUI-based join operation to extract equivalent ICD codes.

The automated mapping was then manually reviewed and refined by (SL, XW) to ensure clinical accuracy. During this refinement process, specific modifications were applied for the 17 shared ADEs identified from FAERS: ‘food craving’ and ‘increased appetite’ were excluded due to a lack of discrete clinical encoding within the ICD code systems. Additionally, ‘nausea’ and ‘vomiting’ were consolidated into a single clinical entity as they share identical ICD codes. This refinement yielded 14 distinct, medically codable ADE categories for longitudinal verification ^1^.

### UT-Physician Dataset ^2^

Serving as the narrow end of our data-driven funnel for targeted clinical validation, the local dataset was derived from the longitudinal UT Physician cohort spanning from January 2015 to June 2025. It contains routine healthcare records in more than 100 clinics across the greater Houston area. The system was served by Allscripts prior to 2020 and subsequently transitioned to EPIC, which includes drug dispensing records reported by more than 50,000 pharmacies across all 50 states and D.C. All clinical records within this system were standardized to the Observational Medical Outcomes Partnership (OMOP) Common Data Model ^35^.

The inclusion criteria required patients to be diagnosed with either: (1) Obesity, identified via specific OMOP condition concept codes; or (2) Overweight with comorbidities, defined as a BMI ≥ 27 kg/m^2^ accompanied by at least one relevant metabolic or cardiovascular comorbidity (e.g., prediabetes, type 2 diabetes, hypertension, obstructive sleep apnea, or fatty liver) documented within one year prior to the measurement. Patients who had undergone obesity-related bariatric surgeries were systematically excluded from the final cohort.

### AOM Exposure Length and Treatment Sessions

We denoted longitudinal treatment sessions as the unit of our temporal dynamic adverse events analysis. Treatment durations were calculated using pharmacy dispensing records from the OMOP *drug_exposure* table, with each continuous session defined by the dispense date plus the documented “days supply”. **Figure 2** illustrates an example of these longitudinal medication trajectories.

**Figure 2.**
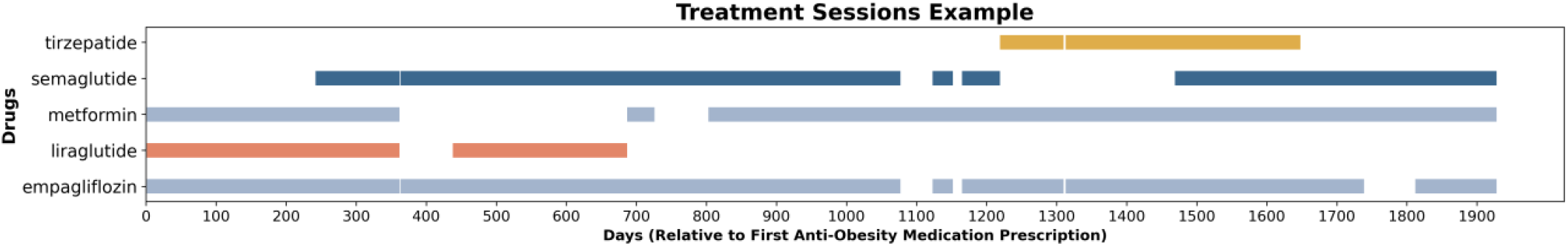
Longitudinal Treatment Sessions Trajectory Example of GLP-1/GIP AOMs

To evaluate how adverse event risks evolve over time, we stratified all sessions into six exposure windows: <30 days, 30–90 days, 90–180 days, 180–365 days (1 yr), 365–730 days (2 yrs), and >730 days. Crucially, to ensure the observed events were incident to the treatment rather than pre-existing, we applied a strict 1-year lookback period. Any targeted adverse conditions recorded within the year prior to a session’s initiation were systematically excluded from the analysis.

### Multivariate Logistic Regression Analysis

We constructed distinct multivariate logistic regression models for each prevalent targeted ADEs validated in the EHR to quantify the dynamic associations between GLP-1/GIP exposure, treatment duration, and ADE risks:

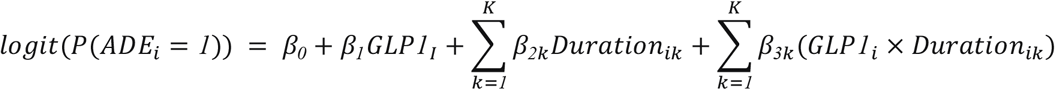

Where *P*(*ADE*_*i*_ = *1*) represents the probability of the adverse event occurrence, *GLP1*_*i*_ is a binary indicator for GLP-1/GIP exposure versus other baseline AOMs, *Duration*_*ik*_ represents the six categorical treatment duration windows. The non-GLP-1 AOM cohort within the initial <30 days exposure window was established as the baseline reference group. Dosage is not considered, as most of the patients are using the standard dosage level.

## Results

### Dataset Characteristics

The baseline characteristics and data volume for both the macro-level FAERS dataset and the micro-level UT-Physician cohort are summarized in **Table 2**. The FAERS dataset comprised 286,461 AOM-related reports, yielding 1,817,760 PT-level adverse event entries. The local EHR cohort incorporated 71,316 patients, accounting for 289,655 distinct treatment sessions.

**Table 2.** Statistics in FAERS and UT-Physician.

|  | **FAERS** | **UT-Physician** |
| --- | --- | --- |
| **Years** | 2014-2025 | 2015-2025 |
| **Reports** | 286,461 | - |
| **Patients** | - | 71,316 |
| **Age, Median (IQR)** | 63, [51,72] | 58, [45,68] |
| **Sex** |  |  |
| **Male** | 126,709 (44.2%) | 26,411 (37.0%) |
| **Female** | 159,021 (55.5%) | 44,890 (62.9%) |
| **Unknown** | 731 (0.3%) | 15 (0.1%) |
| **Drug Exposure** | 388,702 | - |
| **Treatment Sessions** | - | 289,655 |
| **Adverse Event Observation** | 1,817,760 | - |

### FAERS Signal Detection Results

By taking the union of signals detected across all 4 disproportionality algorithms to the FAERS dataset, we captured 754 significant ADE signals for semaglutide, 503 for liraglutide, and 202 for tirzepatide. As illustrated in the Venn diagram (**Figure 3A**), while each medication retained a vast majority of unique signals, intersecting these sets successfully distilled a core profile of 17 shared ADEs consistently associated with the entire drug class.

**Figure 3.**
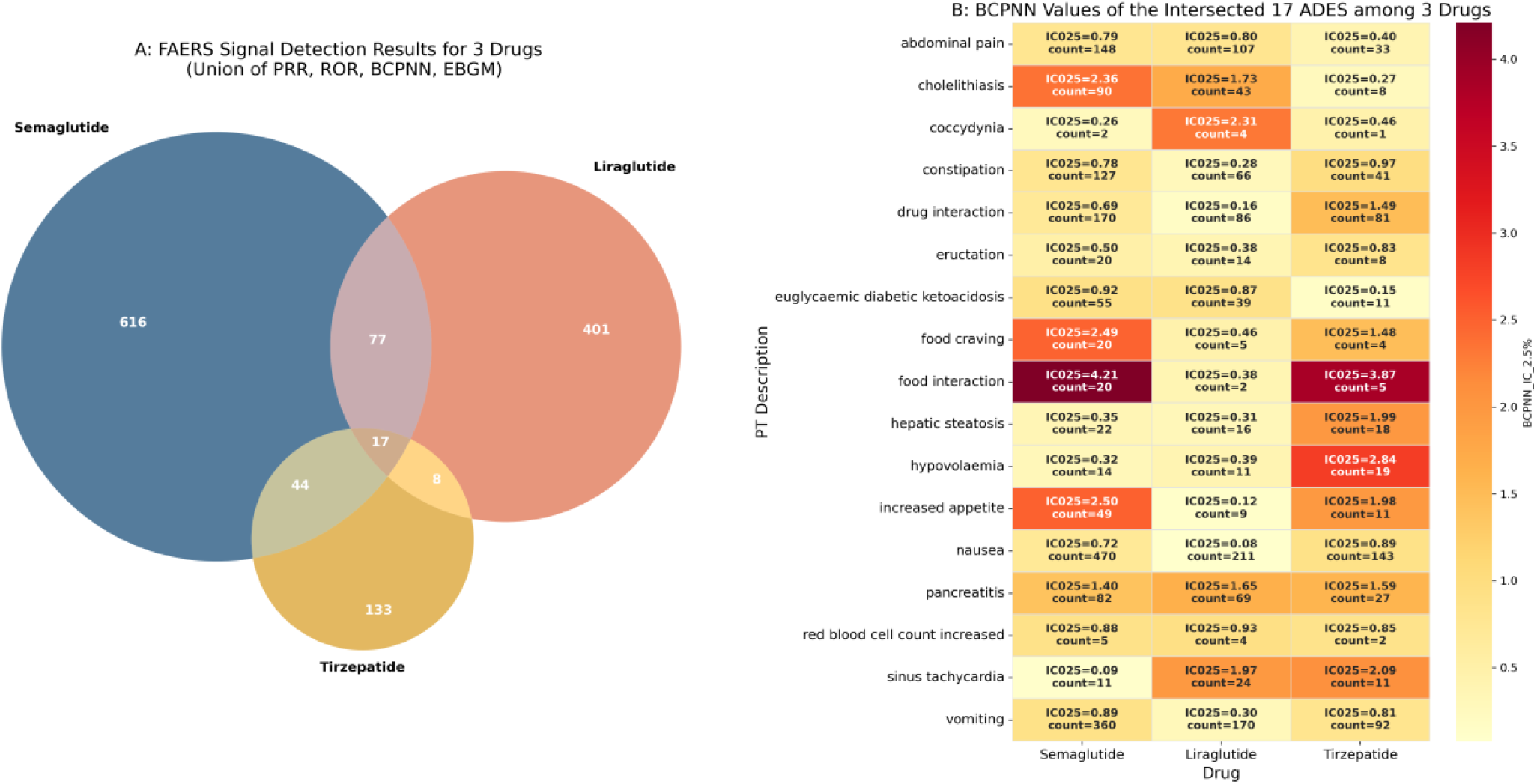
FAERS Signal Detection and Shared ADE Profiles

To evaluate the true signal intensities of these 17 intersected ADEs, we utilized the Information Component (*IC*_*025*_) derived from the BCPNN algorithm as our primary visualization metric (**Figure 3B)**. As a robust Bayesian shrinkage approach, BCPNN effectively stabilizes estimates and minimizes false positives inherent in spontaneous reporting, thereby highlighting prominent class-wide effects alongside distinct drug-specific variations. Significant gastrointestinal and metabolic signals like nausea, vomiting, pancreatitis, and abdominal pain were consistent across all three medications. Furthermore, distinct drug-specific intensities emerged: for instance, semaglutide exhibited strongly elevated signals for cholelithiasis (*IC*_*025*_=2.36) and food craving (*IC*_*025*_=2.49), whereas tirzepatide showed a pronounced signal for hypovolaemia (*IC*_*025*_=2.84). Notably, “food interaction” was captured as an exceptionally strong shared signal across the class (*IC*_*025*_ up to 4.21).

### Treatment Sessions Descriptive Analysis

The distribution of treatment session lengths reveals distinct real-world utilization patterns between GLP-1/GIP therapies and general AOMs (**Figure 4)**. While both cohorts exhibit a right-skewed distribution (**Figure 4B**), GLP-1/GIP RAs usage is significantly more concentrated in short-term intervals. Specifically, the 30–90 day window represents the most frequent session length for GLP-1/GIP RAs therapies (38.8%), with over half (51.6%) of all GLP-1/ GIP RAs sessions terminating within the first 90 days. In contrast, general AOMs demonstrate a more evenly distributed pattern and a notably thicker “long tail” for sustained maintenance. Notably, 12.3% of general AOM sessions extend beyond one year—nearly double the proportion observed for GLP-1/GIP RAs (7.4%).

**Figure 4.**
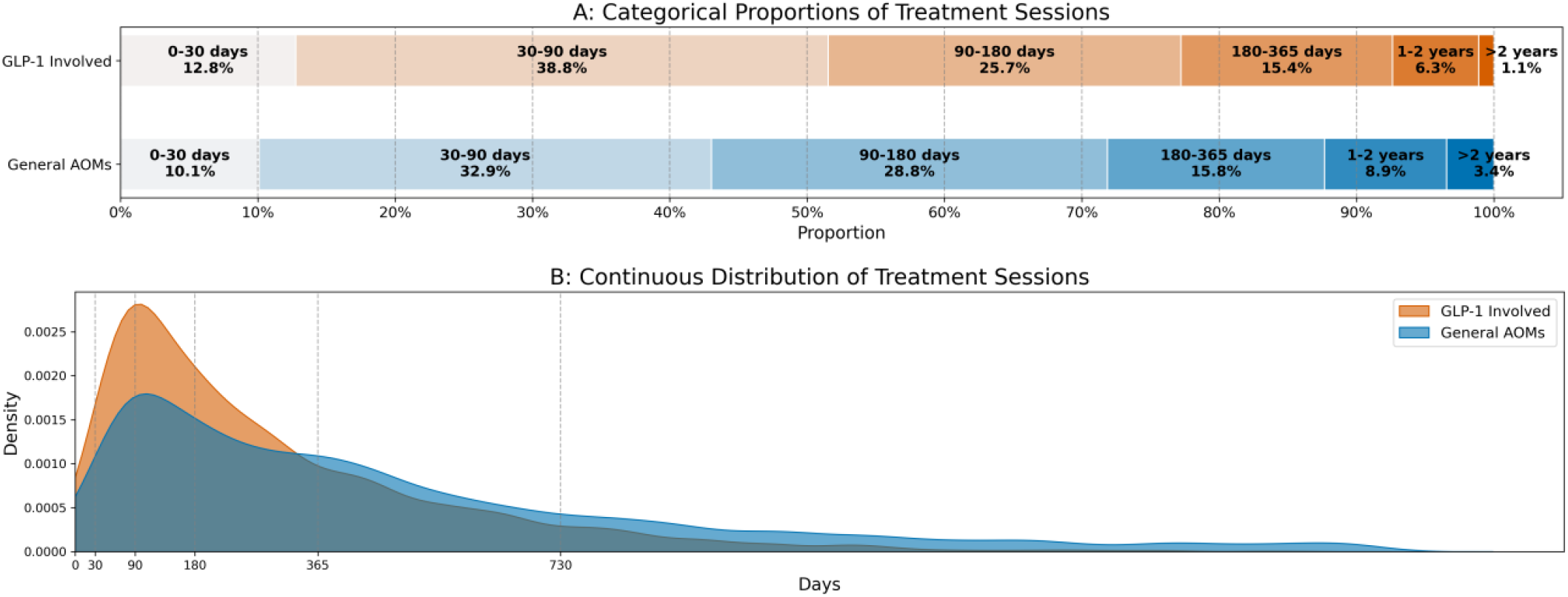
Temporal Distribution of AOM Treatment Sessions

### UT Physician EHR Time-aware Dynamic ADE Signal Results

Following the semantic mapping and rigorous temporal filtering established in our methodology across the refined 14 ADE categories, we identified 141,846 drug-condition pairs with documented timestamps. Within the cohort, 112,885 (79.58%) entries were associated with general AOM therapies, while 28,961 (20.42%) involved GLP-1/GIP RA treatments.

The temporal-stratified disproportionality analysis (**Figure 5**) for the 14 distinct ADEs revealed that absolute case frequencies naturally attenuated over time as treatment prolonged. A majority of the identified signals, particularly gastrointestinal events such as nausea/vomiting and abdominal pain, exhibited their highest PRR and case frequency within the first 90 days of treatment in the UT Physician dataset. Specifically, nausea and vomiting peaked in the 0–30 day window with 598 reported cases. While most signals attenuated as treatment duration increased, the “red blood cell count increased” signal, though the absolute count decreased, demonstrated an upward PRR trend as exposure extended beyond two years.

**Figure 5.**
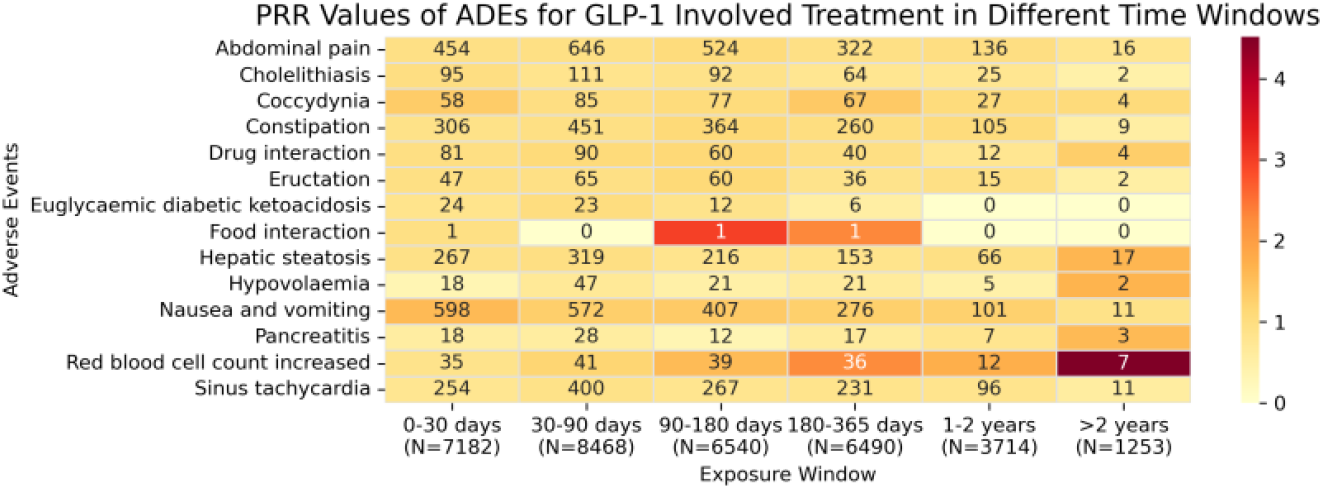
Time-Aware PRR Heatmap of Core ADEs in Local EHR

We then quantified the associations between GLP-1/GIP RAs therapies, session lengths, and ADE occurrences using temporal-stratified multivariate logistic regression (**Figure 6**). Notably, this analysis compares discrete sessions rather than tracking individual patients longitudinally; thus, findings represent cross-sectional associations across duration strata rather than intra-patient causation. Using non-GLP-1 AOMs (<30 days) as the baseline, short-term GLP-1 sessions (0–30 days) were generally associated with lower relative odds for most ADEs (baseline *OR* < *1*.*0*), except for a strong positive association with nausea and vomiting (*OR* = *1*.*9*). Importantly, interaction terms revealed distinct duration-dependent patterns. While initial GLP-1 use correlated highly with nausea and vomiting, longer sessions were associated with comparatively lower odds (interaction *OR* < *1*.*0*). Conversely, extended GLP-1 sessions were positively associated with hepatic steatosis (*OR* > *2*.*5* for >2 years). For cholelithiasis, prolonged GLP-1 exposure demonstrated a negative association, correlating with lower relative odds.

**Figure 6.**
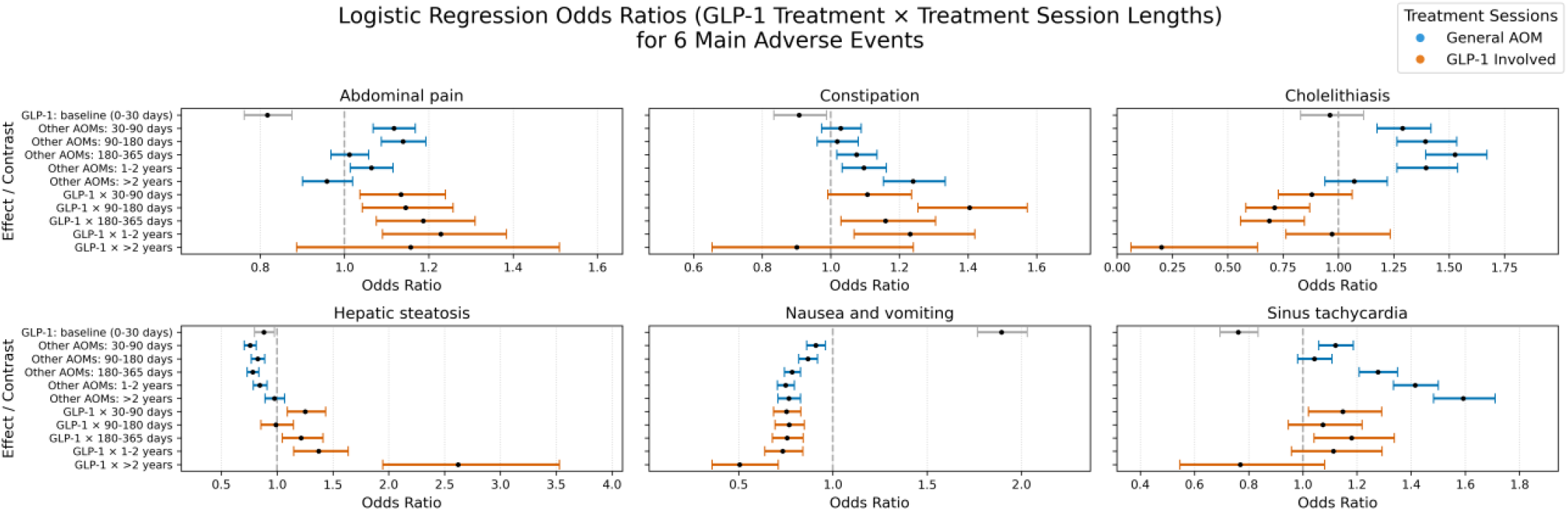
Temporal-Stratified Multivariate Logistic Regression Results

## Discussion

### Findings

In this study, our macro-to-micro pharmacovigilance framework bridged global FAERS reports with local EHR data to systematically evaluate the dynamic safety profiles of GLP-1/GIP RAs. At the macro level, our disproportionality analysis distilled 17 shared adverse events (ADEs). These findings are strongly corroborated by the official drug labels for semaglutide ^36,37^, liraglutide ^38–40^, and tirzepatide ^41,42^, which consistently document prominent gastrointestinal events, including abdominal pain, constipation, nausea, and vomiting. Other physiological conditions are validated in prior literature: pancreatitis has been confirmed across retrospective analyses and clinical trials ^13–15^; cholelithiasis is supported by meta-reviews ^16^; and sinus tachycardia has been observed in both animal models and clinical syntheses ^17–19^. Notably, we also captured an exceptionally strong signal for “food interaction” across the drug class. Rather than representing a traditional toxicological adverse event, we hypothesis this manifests as profound alternations in dietary behaviors, such as early satiety, acquired food aversions, or exacerbated gastrointestinal distress. Thus, the prominent signal may reflect the balance between the GLP-1/GIP RAs therapeutic efficacy and their tolerability challenges.

At the micro level, by stratifying patient data into discrete treatment sessions, we generated new insights into how ADE risks dynamically associate with varying exposure lengths. First, we revealed that GLP-1/GIP RAs therapies are skewed toward short-term use, with over half (51.6%) of treatment sessions terminating within the initial 90 days. This high early-attrition rate was further contextualized by our temporal-stratified models, which demonstrated that the relative odds of nausea and vomiting peak nearly doubled extremely early (0-30 days) but correlate with comparatively lower odds in extended sessions. Similarly, prolonged GLP-1/GIP RAs exposure also demonstrates a negative association with cholelithiasis compared to the initial stages. We hypothesize that these diminishing associations reflect a shared “healthy adherer effect”. Highly susceptible patients who experience severe early gastrointestinal toxicity or early-onset gallstones rapidly discontinue treatment, leaving only tolerant individuals in the long-term cohorts. For cholelithiasis, this pattern further aligns with the recognized kinetics of weight loss: rapid initial weight reduction acts as an acute lithogenic trigger, which naturally diminishes as body weight plateaus during long-term maintenance (> 1–2 years) ^43^.

Our temporal analysis may illuminate previously obscured late-onset risks with entirely distinct trajectories. We identified a positive association between GLP-1/GIP RAs therapies and hepatic steatosis, which significantly increased in extended treatment windows (*OR* > *2*.*5* for >2 years). That is different from recent literature that suggests GLP-1/GIP RAs therapies may attenuate hepatic steatosis ^17,18^. A phase 2 trial ^44^ also suggests that GLP-1/GIP RAs treatment can promote the resolution of nonalcoholic steatohepatitis. We hypothesize that this discrepancy may reflect temporal confounding by indication. It is plausible that patients with progressive or refractory metabolic phenotypes are more likely to insist on continuing GLP-1/GIP RAs therapies due to the drugs’ established metabolic and cardiovascular benefits, such as lowering low-density lipoprotein (LDL), increasing high-density lipoprotein (HDL), and improving overall cardio-renal-metabolic outcomes ^43,45^. Consequently, long-term exposure cohorts might become disproportionately enriched with high-risk individuals undergoing intensive clinical surveillance, naturally leading to a higher rate of incident hepatic steatosis coding over time.

### Limitations

The study has several limitations. First, the FAERS database is inherently subject to underreporting, delayed reporting, and inaccuracies. Additionally, preprocessing steps such as drug normalization and aggregation may introduce minor classification biases. Second, there is a semantic granularity gap in cross-database mapping. MedDRA captures granular, patient-reported symptoms, whereas ICD codes are primarily designed for clinical diagnoses and billing. This fundamental disparity caused certain symptom-level ADEs to drop out while others were consolidated (e.g., merging “nausea” and “vomiting”). Third, although restricting our local EHR analysis to the 17 core ADEs shared across all three GLP-1/GIP RAs successfully highlighted class-wide effects, it inevitably sacrificed unique, drug-specific safety signals. Fourth, our analysis relies on a single health system (UT-Physician), which limits generalizability and introduces a duration-dependent observation bias. As the treatment window extends (e.g., 1-2+ years), uncaptured out-of-network care, telehealth usage, and out-of-pocket purchases disproportionately underestimate late-onset ADEs. Fifth, our temporal models did not explicitly adjust for dosage titration. As clinical protocols mandate gradual dose escalation for GLP-1/GIP RAs, patients in extended exposure windows are inherently more likely to be receiving higher maintenance doses. Thus it’s challenging to disentangle cumulative duration effects from the higher maintenance doses inherent to extended therapies. Finally, our identification of conditions was based on defined “treatment sessions” involving GLP-1/GIP RAs therapies. However, these sessions did not strictly isolate GLP-1/GIP RAs monotherapy. Patients often present with complex polypharmacy or other treatments at the same time. We cannot fully exclude the influence of concurrent background medications, procedures, or drug-drug interactions. Consequently, while our temporal-stratified models identified strong dynamic associations, they inherently cannot establish definitive causal links between drug usage and the observed ADEs.

## Conclusions

In this study, we proposed a macro-to-micro pharmacovigilance framework to systematically evaluate the dynamic safety profiles of GLP-1/GIP RAs. By bridging global FAERS signal detection with local, session-based EHR data, we distilled a core profile of 17 shared adverse events (ADEs) that robustly corroborate existing clinical trials and literature. Furthermore, our time-aware analysis demonstrates that GLP-1/GIP RAs safety profiles are not static; most of them are profoundly duration-dependent. Traditional methodologies often lack the extended follow-up required to track multi-year medication trajectories. Our localized, temporal-stratified approach provides a uniquely powerful paradigm that addresses this gap, offering dynamic insights and generating actionable clinical hypotheses for safer, more personalized long-term obesity management.

## Data Availability

All data produced in the present study are available upon reasonable request to the authors

## Acknowledgements

This work was funded by the National Human Genome Research Institute (3R01HG012748-02S2), Cancer Prevention and Research Institute of Texas Established Investigator Award (RR230020), and the National Institute of Aging (R01AG072799-03).

## Author Contributions

Lu, S conceived the study under advice of Ruan, X. Lu, S designed and implemented the programs, performed data analysis, and drafted the manuscript. Wang, L contributed to data processing. Wang, X participated in study design and literature review. All authors contributed to the writing and editing of the manuscript.

## Footnotes

1 The mapping can be accessed through: https://github.com/Frank-LSY/GLP-1-Temporal-Drug-Adeverse-Event-Resource.

2 The University of Texas Institutional Review Board (IRB) approved the study, and the Ethics Committee waived written consent from participants.

